# Self-injury, suicidal ideation and -attempt and eating disorders in young people following the initial and second COVID-19 lockdown

**DOI:** 10.1101/2022.03.08.22271980

**Authors:** Stine Danielsen, Andrea Joensen, Per Kragh Andersen, Trine Madsen, Katrine Strandberg-Larsen

**Affiliations:** Danish Research Institute for Suicide Prevention – DRISP, Mental Health Center Copenhagen, Postal address: Kildegaardsvej 28, Opg. 15, 4. floor, DK – 2900 Hellerup; Section of Epidemiology, Faculty of Health and Medical Sciences, University of Copenhagen, Postal address: Øster Farimagsgade 5, bd. 24, PO Box 2099, DK - 1014 Copenhagen K; Section of Biostatistics, University of Copenhagen., Postal address: Øster Farimagsgade 5, bd. 5, PO Box 2099, DK - 1014 Copenhagen K; Section of Epidemiology, University of Copenhagen, Faculty of Health and Medical Sciences., Postal address: Øster Farimagsgade 5, bd. 24, PO Box 2099, DK - 1014 Copenhagen K

**Keywords:** COVID-19, lockdown, self-injury, suicidal behaviour, eating disorder, youth, longitudinal data, repeated cross-sectional data

## Abstract

**Background:** The initial COVID-19 lockdowns have had negative effect on different mental health measures, especially in young women. However, the impact on self-injury, suicidality and eating disorder (ED) are less elucidated and remains inconsistent. We compare self-reported self-injury, suicide ideation and -attempt and symptoms of EDs from before through different pandemic periods until spring 2021.

**Methods:** Young participants in the Danish National Birth Cohort reported these measures in an 18-year follow-up in 2015-2021 and in a COVID-19 survey in spring 2021 when participants were aged 19-24 years. Changes in measures from pre to post lockdown were estimated with longitudinal data (N=7,597) and with repeated cross-sectional data (N=24,625) by linear regression.

**Findings:** In the longitudinal comparisons 14% of women and 7% of men reported self-injury pre lockdown, which decreased 6%-points (95% CI:-7%;-5%) for women and 3%-points (95% CI:-4%;-2%) for men during lockdown. For suicide ideation, the pre lockdown proportions were 25% and 18% for women and men respectively, and decreased 7%-points (95% CI:-8%;-6%) for women and 3%-points (95% CI:-5%;-1%) for men. For suicide attempt no change was observed. Pre lockdown 15% and 3% of women and men, respectively, had symptoms of EDs, which decreased 2%-points (95% CI:-3%;-1%) for women. We observed no changes in proportions of self-injury, suicide ideation or EDs in the repeated cross-sectional data.

**Interpretation:** Our findings provide no support for increase in self-injury, suicidality and EDs following the lockdowns, and if anything, indicate a reduction in self-injury and suicide ideation as well as EDs in women.

## Introduction

In the beginning of 2020 COVID-19 quickly spread globally and was on 11^th^ March declared a global pandemic by the World Health Organization which led to public health measures being implemented to mitigate the spread of COVID-19. Young people are at low risk of being severely ill by COVID-19 but have been suggested as most vulnerable to the collateral damages of lockdown and several studies have found an aggravation in the mental health among young people during the initial lockdown, especially among young women^1-5^. However, literature exploring whether this aggravation in mental health also has manifested as changes in self-injury, suicidality and eating disorders (EDs) during lockdown is sparse and with inconsistent findings. The majority of pre to post lockdown comparisons of self-injury and suicidality are based on health registries, which are compromised by the general reduction in patient contacts during lockdown and mostly refers to registrations of cases brought to hospital due to somatic injuries^6^. A Danish register-based study found a signal for increase in hospital registered suicidal behaviour in young people aged 18-29 years during the first lockdown, but no change during the entire first year following the lockdown^7^. Register-based studies from other countries have documented a decrease in self-injury and suicide attempt among young people^8-11^. Studies from Norway and Korea based on self-reported repeated cross-sectional data, found no pre to post lockdown change in suicide ideation and a decrease in suicide ideation and -attempts respectively^12-13^. Contrary, a longitudinal study based on self-reported data in China demonstrated an increase in self-injury and suicide ideations and -attempts during spring 2020^14^. An international register-based study including pre-liminary suicide data from 21 countries in all age groups, showed no evidence of increased suicide rates during the first year of the pandemic^15^. However, other studies have suggested increased suicide rates during lockdown when including young people only and pre-liminary and unvalidated data based on Danish registers reviled a signal of an increased number of suicides among young women aged 20-24 years^16-18^.

Regarding EDs, studies from USA and Canada found that hospital admissions and new diagnosis for restrictive EDs were twice as high among adolescents during the first year of lockdown compared to previous years^19-21^. To our knowledge, no studies have compared pre- and post-pandemic self-reported data on EDs among young people.

To mitigate the spread of COVID-19, the Danish government, similar to many other countries, implemented a national lockdown in March 2020 requiring a closure of schools, daycare centers, sport facilities, restaurants, shops etc. and working from home was either mandatory or highly recommended in non-critical functions^22^. The restrictions were slowly lifted during spring 2020, but gradually reinforced during fall, and in December a 2^nd^ national lockdown was declared. This 2^nd^ lockdown turned out to be more prolonged and was slowly lifted during spring 2021.

### Aim

In this study we compare the proportion of young Danish people reporting self-injury, suicide ideation and –attempts and symptoms of EDs with similar pre and post lockdown data across the two national lockdowns. Further, we examined if the lockdown disproportionally impacted men and women.

## Materials and methods

### The Danish National Birth Cohort

The Danish National Birth Cohort (DNBC) is a nationwide cohort established in the mid-nineties, which include about 30% of all children born in Denmark in 1996-2003^23^. Data from prenatal life through early adulthood has been collected with the latest collection being the 18-year follow-up (DNBC-18). The DNBC-18 data collection started in 2016 and was completed ultimo 2021 when the last participant reached 18 years and three months, which was the age of invitation. Further information about the cohort and the DNBC-18 is available at www.dnbc.dk. For our purpose, we used two different study populations; one with longitudinal data collected pre and post lockdown, and one consisting of repeated cross-sections of lockdown appropriate periods in 2020-2021, as well as similar periods in 2018-2019, Figure 1. Both study populations were restricted to DNBC participants with information on the baseline covariates household-socio-occupational status, maternal age at childbirth, parity and maternal smoking collected during pregnancy in order to create sampling weights.

**Figure 1.**
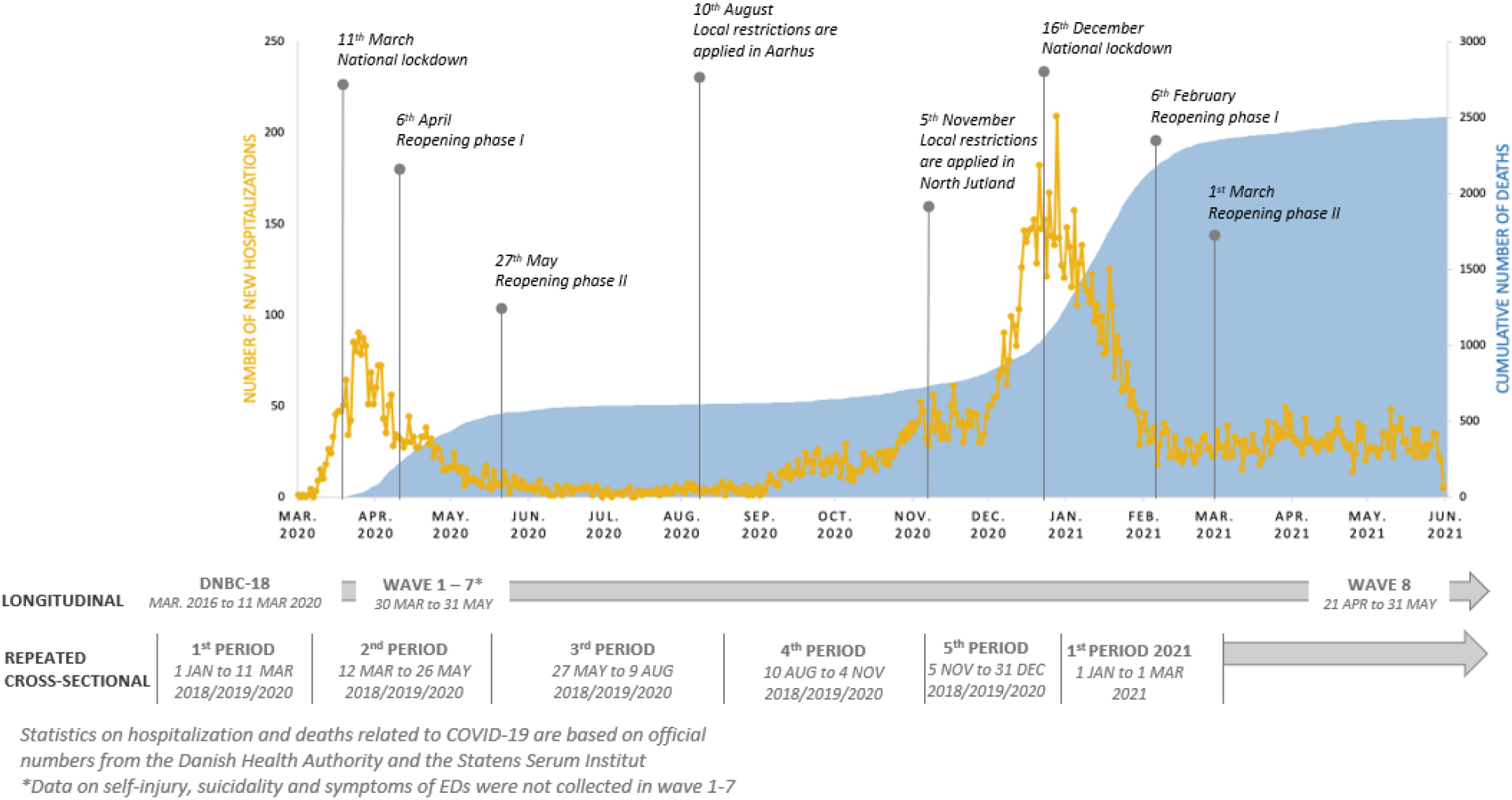
The development of the COVID-19 pandemic, aligned with seminal events during the lockdown in Denmark, together with the timing of the data collections

### Longitudinal data

In April 2020 during the initial lockdown, a COVID-19 survey was launched and consisted of 7 weekly waves^22^. In April-May 2021 when the second national COVID-19 lockdown was gradually lifted, the DNBC invited participants to complete an additional wave, i.e. wave 8. All DNBC participants with an active social security number, who had not actively withdrawn their participation, and provided either their private mail or phone number, were invited. The population in this study was restricted to 27,441 participants who completed the DNBC-18 prior to 11^th^ March 2020, and provided complete data on self-injury, suicidality and EDs, Figure S1. Of these, 7,597 participants also completed wave 8 of the COVID-19 survey.

### Repeated cross-sectional data

Participants in the DNBC were born over a period spanning 8 years. Thus, per design, they completed the DNBC-18 in different years, and we exploited this feature to perform cross-sectional comparisons of participants completing the DNBC-18 between 1^st^ January 2018 and 11^th^ March 2020 with those completing hereafter until 1^st^ March 2021. This study population includes 24,625 participants sub-divided into year 2018-2021, Figure S2. Based on the date completing the DNBC-18, the participants were assigned to one out of 16 different periods; 11 periods representing pre lockdown (January 2018-11^th^ March 2020) and 5 periods representing post lockdown (12^th^ March 2020-1^st^ March 2021), Figure 1.

### Measures of self-injury, suicidality and EDs

To measure self-injury, suicide ideation and suicide attempt, two items in the DNBC-18 and one item in the 8^th^ wave COVID-19 survey were used to measure whether these behaviours had occurred within the last year (yes vs. no), Table S1. Self-injury was worded as “have you harmed or hurt yourself on purpose within the last year” and suicide ideation was worded as “have you thought about taking your own life (even though you would not do it) within the last year”. Suicide attempt was worded as “have you tried to take your own life within the last year”. Symptoms of EDs were collected in the DNBC-18 with items adapted from the McKnight Risk Factor survey on weight and shape concerns and items from the Youth Risk Behavior Surveillance System survey on binge eating, self-induced vomiting, and use of laxatives^24,25^. We defined EDs in accordance with definitions used and described by Micali et al^26^ to classify threshold and sub-threshold anorexia, bulimia, purging disorder, and binge eating disorder, Table S2 and Table S3. Because of the low frequency of threshold EDs in men we chose to combine threshold and sub-threshold EDs into one measure reflecting symptoms of EDs within the last month (yes vs. no). Analyses of threshold EDs with no gender-stratification are included in the supplement.

## Statistical analysis

Sampling weights were estimated to account for differential attrition in DNBC-18 and wave 8 of the COVID-19 survey. For the inverse probability weighting (IPW) we used logistic regressions with participation, i.e. having data as outcome and the following predictors: gender, household-socio-occupational status, maternal age at childbirth, parity, and maternal smoking collected during pregnancy, categorized as shown in Table S4. Separate analyses were performed for DNBC-18 and wave 8 of the COVID-19 survey and performed on the relevant baseline populations described in Figure S1 and S2. The relevant sampling weights were used in all analyses in this study.

Gender-specific proportions of self-injury, suicidality and symptoms of EDs were estimated with corresponding 95% confidence intervals (CI) pre (DNBC-18) and post (wave 8) lockdown, with the longitudinal data, and for each period in the repeated cross sections to illustrate any differences. Subsequent, we estimated changes in the proportion of young people with self-injury, suicide ideation and -attempt as well as symptoms of EDs from pre to post lockdown by fixed effect linear regression on the longitudinal data and linear regression on the cross-sectional data. We used linear regressions, instead of logistic regressions in order to get an estimate of the absolute change (i.e., in percentage-points) of the proportions reporting self-injury, suicidality, and symptoms of EDs from pre to post lockdown. Separate regressions were conducted for self-injury, suicide ideation and - attempts, and symptoms of EDs respectively. To examine for disproportional impact of the lockdown on any of the outcomes in men and women, we added an interaction between lockdown (pre vs. post) and gender. In the linear regressions on the repeated cross-sectional data, we initially tested if the impact of lockdown varied across periods by including an interaction between lockdown (pre vs. post) and period, and if insignificant this interaction was omitted.

The results in the longitudinal setup may be biased as the participants all were 18-years at the pre lockdown measure but between 19 and 24 years at the post lockdown measure. In sensitivity analyses we addressed this by restricting to participants aged 19-20 years when completing wave 8 to limit the time-gap. Further, we did sensitivity analyses in the repeated cross-sectional data where self-injury and suicide ideation were restricted to being within six months or four weeks instead of one year. All analyses were performed unadjusted and with SAS Software, version 9.4 (SAS Institute, North Carolina, US) and the level of statistical significance was *P* <.05.

## Results

### Longitudinal data

Our study population were aged 19-23 years with a median age of 20·9 years in spring 2021 and more women than men responded in the DNBC-18 and wave 8. Less than 10% were living without parents and 5% had another occupation than school, Table S4. In the analyses of the longitudinal data, the proportion of self-injury before lockdown was 14% and 7% among women and men, respectively, and decreased post lockdown in both women and men, Figure 2a. The proportion reporting self-injury decreased with 6%-points (95% CI:-7%;-5%) among women and 3%-points (95% CI:-4%;-2%) among men, Figure 3a. For suicide ideation, the absolute decrease in percent-points was similar, as it was 7%-points (95% CI:-8%;-6%) in women and 3%-points (95% CI:-5%;-1%) in men, but the pre lockdown proportion of suicide ideation were 25% and 18% respectively, Figure 2b and 3b. Thus, the reductions were smaller relatively. The pre lockdown proportion of suicide attempt was 0·9% in women and 0·5% in men and the proportion did not change significantly during lockdown for women or men, Figure 2c, Figure 3c. Prior to lockdown 15% of women and 3% of men reported symptoms of EDs within the last month, Figure 2d. Post lockdown these proportions decreased 3%-points (95% CI:-4%;-2%) among women, and 1%-points (95% CI:-2%;0%) among men, Figure 3d.

**Figure 2.**
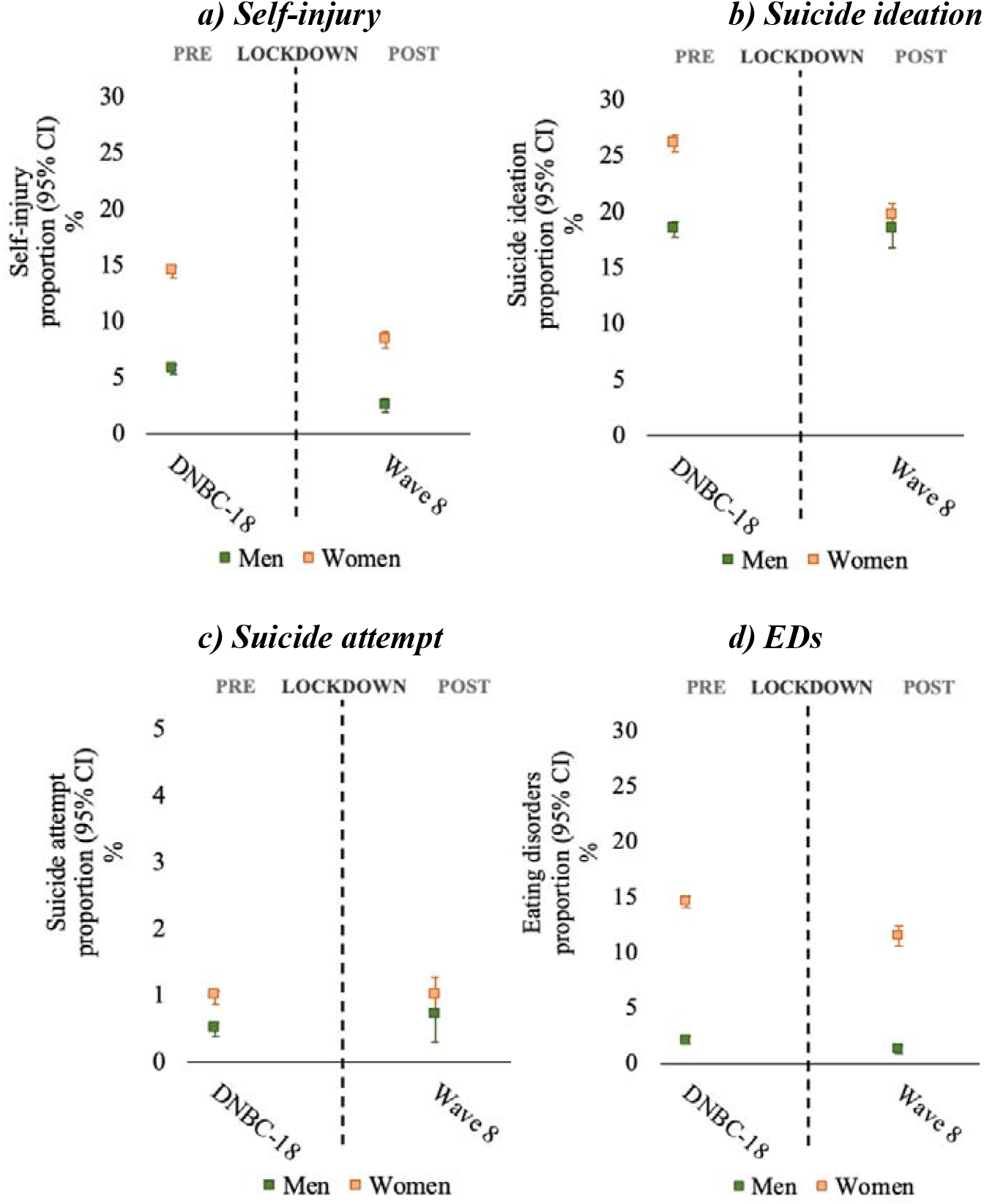
Proportion of self-injury, suicide ideation and attempt, and EDs pre and post lockdown among men and women based on the longitudinal data collected in the DNBC-18 (N=27,441) and wave 8 (N=7,597) of the COVID-19 survey, approximately one year post the initial lockdown.

**Figure 3.**
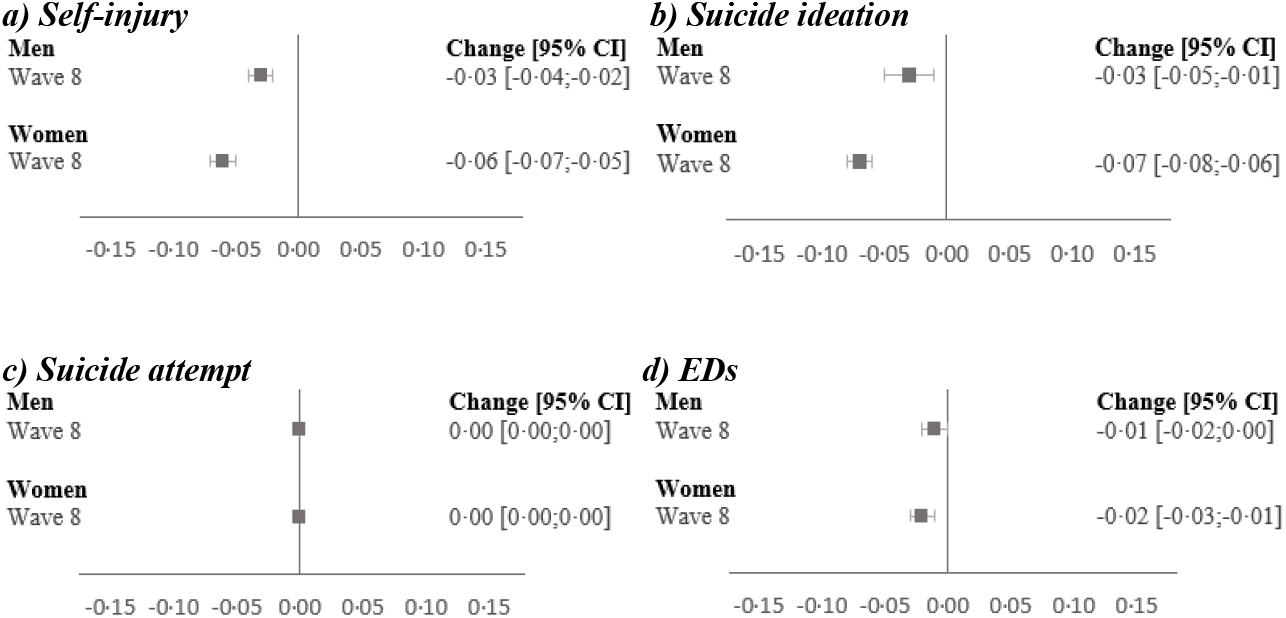
Change in the proportion of self-injury, suicide ideation and attempt and EDs from pre to during lockdown in men and women based on the longitudinal data collected in the DNBC-18 (N=7,597) and wave 8 of the COVID-19 survey (N=7,597), approximately one year post the initial lockdown.

Our sensitivity analyses restricted to participants aged 19-20 years when completing wave 8, i.e. completing the DNBC-18 in 2019 or early 2020, resulted in similar results for suicide ideation and - attempt and symptoms of EDs following lockdown, while the decrease in self-injury were slightly smaller, Figure S3. Further self-injury and suicide ideation restricted to being within six months or four weeks instead of one year in the repeated cross-sectional data did not change the results (data not shown).

### Repeated cross-sectional data

In the analyses of the repeated cross-sectional data, women had higher proportions of self-injury, suicide ideation and symptoms of EDs than men. The calendar periods showed no clear pattern of self-injury, suicide ideation and -attempt, and symptoms of EDs and there was no year-to-year variation of the measures, Figure 4. The interaction analyses showed no statistically significant change in the proportions of any outcomes from pre to post lockdown by neither gender nor by any of the calendar periods, Figure 5.

**Figure 4.**
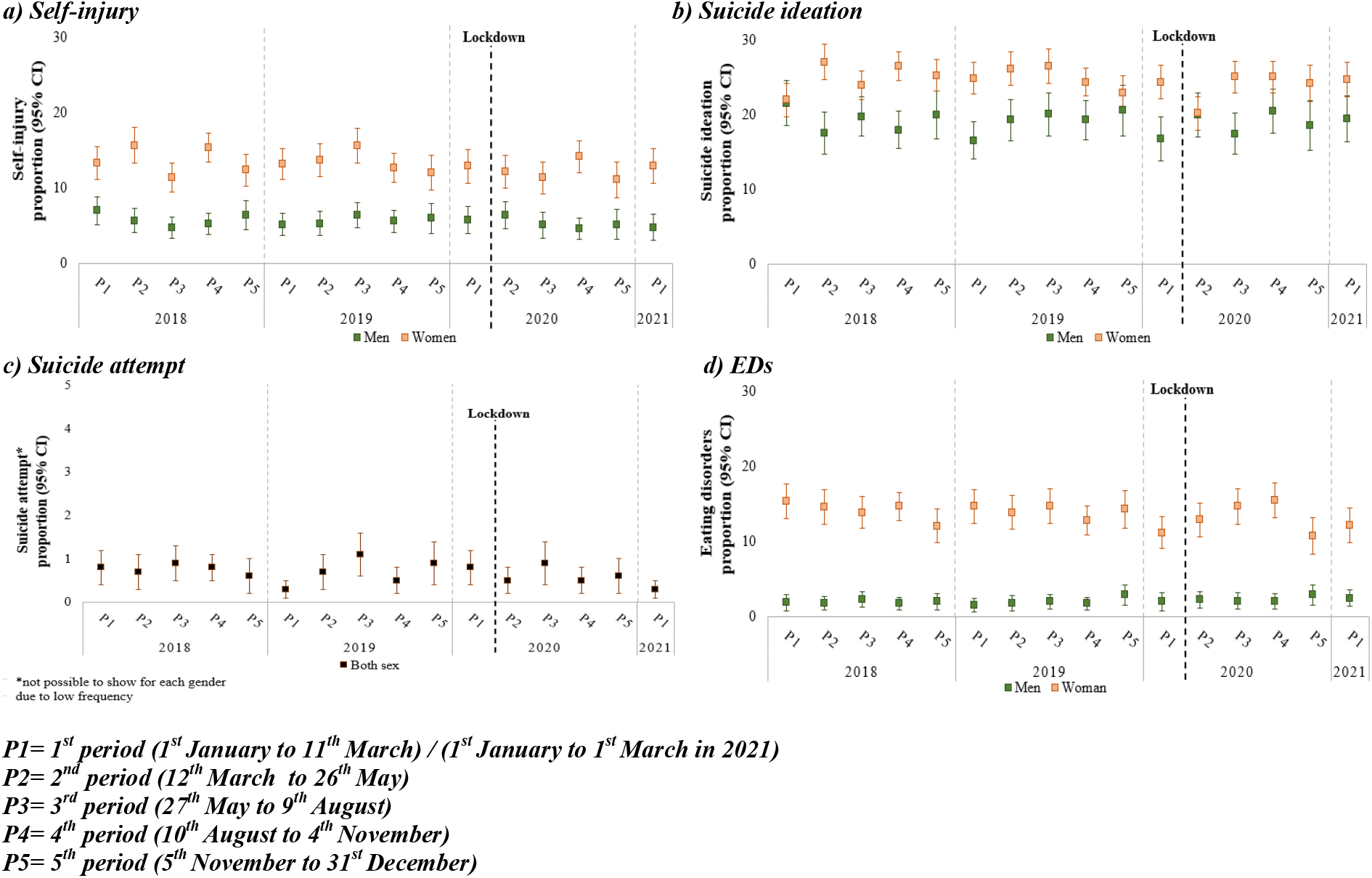
Proportion of self-injury, suicide ideation and -attempt, and EDs pre and post lockdown in men and women based on repeated cross-sectional data collected in the DNBC-18 (N=24,625) in 2018-2021.

**Figure 5.**
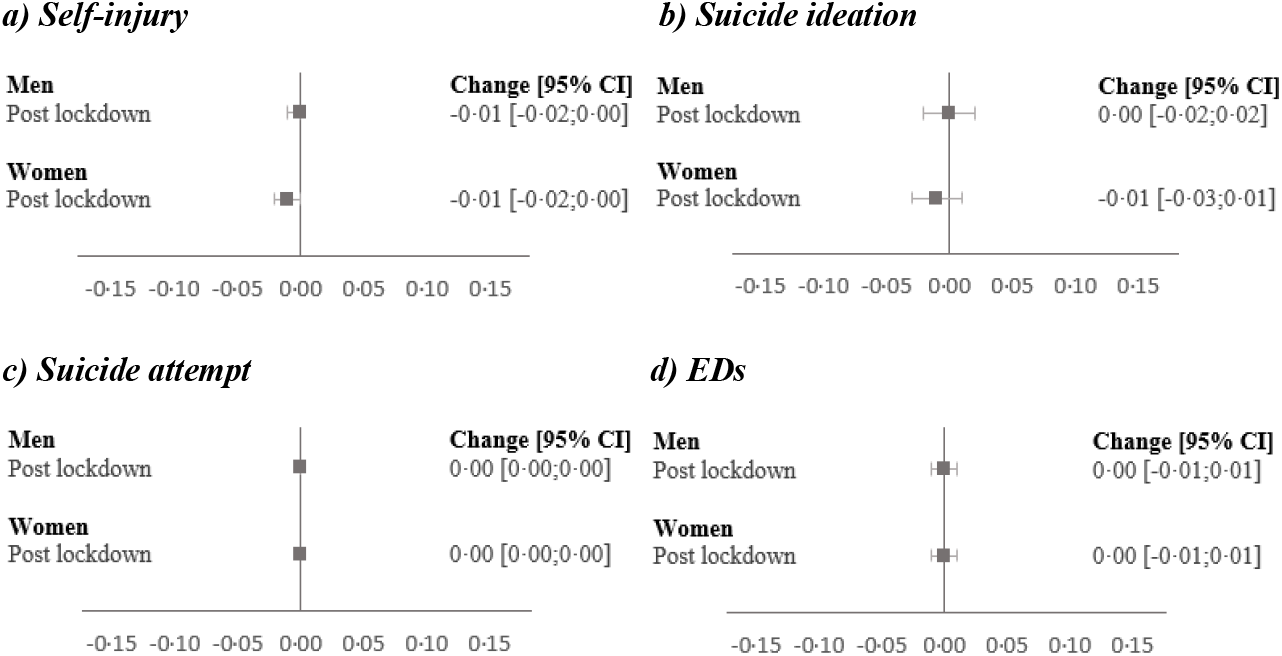
Change in the proportion of self-injury, suicide ideation and attempt and EDs from pre to during lockdown in men and women based on the repeated cross-sectional data collected in the DNBC-18 (N=24,625) in 2018-2021.

## Discussion

In this study with tandem use of longitudinal and repeated cross-sectional data, we observe that the lockdowns, including a more prolonged second lockdown during the winter season, resulted in no increase in self-injury, suicide ideation and -attempt, or symptoms of EDs. If anything, our longitudinal data, indicate a post lockdown reduction in self-injury among both men and women, and smaller reductions relatively for suicide ideation and EDs.

Thus, our findings do not support that the aggravation in mental health documented in several countries following lockdown, have yet resulted in young people having a higher risk of self-injury, suicidality, and EDs. As outlined in the introduction, findings from previous studies are inconsistent. The mixed findings in the literature may result from methodological differences. Cross-sectional studies using health register data all found a decline in self-injury and suicidal behaviour related hospital contact during lockdown^7-11^. This reduction is attributable to bias caused by the general reduction in health care use and it is important to note that self-injury and suicidality with hospital contact only covers the most severe cases. However, cross-sectional studies using self-reported data, including this study, also found either a reduction or no change in self-injury and suicide ideation and -attempts during lockdown^12-13^. In contrast, a longitudinal study in China with self-reported data have suggested an increase in both self-injury and suicide ideation and -attempts during lockdown^14^. The longitudinal study only included four months of lockdown, had a small population (n=1,241) and participants were only aged 9-16 years which could, together with differences in the lockdowns, explain why the findings differ from ours even though the methodology is very similar. We were not able to include suicides from the cause-of-death register to explore the potential increase in suicide among especially young women as suggested by preliminary unvalidated Danish data and current international literature^15-18^. Regarding EDs, previous studies used data on new ED diagnosis or hospital admissions which increased during lockdown contrary to our findings^19-21^. As there was a general reduction in health care use due to lockdown, this increase may even be understated but differences in health care systems and thereby the registers may also affect the comparability between studies. Further, these health register measures of EDs cover more severe cases compared to our self-reported symptoms of EDs. However, our sensitivity analyses of symptoms that fulfilled our definition of threshold EDs (weekly symptoms) likewise indicated no signal of an increase following lockdown, Figure S4.

In general, the previous literature has varied with regard to follow-up time, origins from different countries, the course of the pandemic, the public health precautions, e.g. the extend of lockdown and thus the impact on self-injury, suicidality, and EDs may be different. Further, even small age differences in study populations focusing on young people may explain the inconsistent result. Importantly our population mainly consisted of young people studying and still living at their parental home and cannot necessarily be generalized to older populations who are more on their own and therefore could be more vulnerable to the collateral damages of lockdown.

Possible reasons for the decrease in self-injury, suicidality, and EDs in the longitudinal data is that the social distancing actually has been beneficial for some young people^27^. Being closer to their families, having more time and less obligations to activities such as sports, part-time jobs and parties, i.e. generally a reduction in the pressure of living up to the social norms for young people could have impacted the mental health in a positive direction. Further, it is possible that the impact of lockdown has been positive in some groups while negative in other groups which equalizes the impact. A similar DNBC study found slight interim deterioration in mental health in young people without pre-existing depressive symptoms following lockdown while no differences were observed in young people with pre-existing depressive symptoms^1^. Furthermore, the initial negative impact on mental health quickly attenuated and may have been a shock effect that did not manifest as self-injury, suicidality, and EDs.

## Supporting information

Supplemental Tables and Figues

## Data Availability

According to European law (General Data Protection Regulation), data containing potentially identifying or sensitive personal information are restricted. However, for academic researcher, data could be available on request via DNBC

## Strength and limitations

This study is the first to compare proportions of self-injury, suicidality, and symptoms of EDs in different periods during the first year of lockdown using both longitudinal and repeated cross-sectional data from a large population. A major strength of this study is the use of self-reported data that captures more subtle cases than register data and still provides relevant information for screening and prevention purposes^28,29^. Further our study populations are from a large cohort consisting of relatively healthy and well-functioning young people.

The results using longitudinal and repeated cross-sectional data were not completely consistent. Although both suggested that the proportion of young people with self-injury, suicidality, and EDs did not increase during the lockdown. Different strengths and limitations in the study designs could have resulted in the different results; 1. As the longitudinal data includes a specific COVID-19 related survey, the participants may have over- or understated their answers in accordance with their feelings of lockdown making the comparison with the pre-lockdown measure biased. This is less of a concern in the repeated cross-sectional data only using the DNBC-18, as it was an ongoing survey that did not mention the COVID-19 pandemic in any way; 2. When using longitudinal data there is a risk of differential attrition as young people with mental health problems are less likely to participate in follow-ups^30^. This may have resulted in bias as the observed decline in self-injury, suicide ideation, and EDs in wave 8 may be explained by loss to follow-up rather than the lockdown, however we used sampling weight to limit this bias. The repeated cross-sectional data is only vulnerable to attrition if the participation in DNBC-18 systematically changed over year of birth, which does not seem plausible; 3. A major strength of the longitudinal data is that we analyse the same young people pre and post lockdown and thereby all time-invariant factors are adjusted for. In the repeated cross-sectional data, we compared groups of young people based on birth year and the results may reflect factors related to birth year rather than lockdown. However, the level of self-injury, suicidality, and symptoms of EDs were stable during the entire pre lockdown period and thus it is unlikely that a sudden change would have happened in the absence of lockdown. 4. A strength in comparing different young people aged 18 years pre and post lockdown is that the age is adjusted for. In the longitudinal data, the pre lockdown measures were collected at age 18 while the post lockdown measures were collected at age 19-23. Thus, the time interval between the pre- and post-measure varied and as many major events such as graduating high school, moving away from parents and starting to shape the future happens in this age range the results may be biased. However, sensitivity analyses showed similar results when restricting the time interval to two years indicating that the varying time interval cannot explain the entire observed decrease in self-injury, suicide ideation and symptoms of EDs during lockdown; 5. The measures of self-injury and suicidality were defined as being within the last year. Thus, in the longitudinal setup the measures were restricted to an entire year of lockdown and thereby adjusted for seasonal differences even though the pre lockdown data were collected at different times of the year. In the repeated cross-sectional data, the post-lockdown measures will somewhat overlap the pre-lockdown period until the last lockdown period that covers an entire year of lockdown. However, analyses where self-injury and suicide ideation were restricted to being within six months or four weeks instead of one year did not change the results. Symptoms of EDs were defined as being within the last month. Seasonal differences in EDs could bias the results in the longitudinal data as the pre and post lockdown data mainly were collected at different calendar periods. However, the repeated cross sections did not reveal any clear seasonality, as the pre-defined lockdown relevant periods represent different periods of the year.

In conclusion, this study suggests that the lockdown, did not result in increased proportions of young people with self-injury, suicidality, or symptoms of EDs. Findings from longitudinal analyses even indicate that the proportion of self-injury and suicide ideation has decreased slightly post the lockdowns in both men and women, while a minor decrease in symptoms of EDs were observed only in women.

## Funding

The Velux Foundation

## Author contributions

SD, AJ, PKA, and KSL conceived and designed the study. AJ, KSL, and SD were involved in the data collection and data management of the DNBC-18 and COVID-19 survey. SD conducted the analyses, supervised by AJ and PKA. SD and KSL drafted the first draft of the manuscript. TM contributed with specific inputs for the suicide behavior categorization. All authors contributed to the analytical approach and interpretation of the data, revisions of the manuscript, and submission of the final manuscript. The corresponding author attests that all listed authors meet authorship criteria and that no others meeting the criteria have been omitted and is the guarantor of the manuscript.

## Declaration of interest

All authors declare no competing interests.

## Data sharing

According to European law (General Data Protection Regulation), data containing potentially identifying or sensitive personal information are restricted. However, for academic researcher, data can subsequent to approval be available. However, any request for data needs to follow the outlined procedures, see: https://www.dnbc.dk/access-to-dnbc-data

## Funding

This study was made possible by a grant from the Velux Foundation (grant number 36336, ‘Standing together at a distance – how Danes are handling the corona crisis’) The funders of the study had no part in the conception or design of the study, or in the decision to publish.

## Ethics

This study was approved by the Danish Data Protection Agency via a joint notification to the Faculty of Medicine and Health Sciences – University of Copenhagen (ref. 514-0497/20-3000, ‘Standing together at a distance: how are Danish National Birth Cohort participants experiencing the corona crisis?’). The cohort is approved by the Danish Data Protection Agency and the Committee on Health Research Ethics under case no. (KF) 01-471/94. Data handling in the DNBC has been approved by Statens Serum Institut (SSI) under ref. no 18/04608 and is covered by the general approval (Fællesanmeldelse) given to SSI. The 18-year follow-up was approved under ref. no 2015-41-3961. The DNBC participants were enrolled by informed consent.

## Acknowledgment

The Danish National Birth Cohort (DNBC) was established with a significant grant from the Danish National Research Foundation. Additional support was obtained from the Danish Regional Committees, the Pharmacy Foundation, the Egmont Foundation, the March of Dimes Birth Defects Foundation, the Health Foundation, and other minor grants. The DNBC Biobank has been supported by the Novo Nordisk Foundation and the Lundbeck Foundation. Follow □ up of mothers and children has been supported by the Danish Medical Research Council (SSVF 0646, 271□08□0839/06□066023, O602□01042B, 0602□02738B), the Lundbeck Foundation (195/04, R100□A9193), The Innovation Fund Denmark 0603□00294B (09□067124), the Nordea Foundation (02□2013□2014), Aarhus Ideas (AU R9□A959□13□S804), a University of Copenhagen Strategic Grant (IFSV 2012) and the Danish Council for Independent Research (DFF – 4183□00594 and DFF – 4183□00152). The 18-year follow-up specifically was funded by the Danish Council for Independent Research (DFF-4183-00594B (Close to Adult: 17-year follow-up of the Danish National Birth Cohort)).

## Notes

### Competing Interest Statement

The authors have declared no competing interest.

### Author Declarations

The Research Ethics Committee for SCIENCE and HEALTH (University of Copenhagen) finds, according to information received, that the project is compliant with relevant Danish and International standards and guidelines for research ethics.

