## Supplemental Tables and Figues for "Self-injury, suicidal ideation and -attempt and eating disorders in young people following the initial and second COVID-19 lockdown"

**Supplement**

**Table S1.** Items on self-injury and suicidality in the DNBC-18 and wave 8 of the COVID-19 survey

| **Questions in the DNBC** | **Response options** | **Variable coded as** |
| --- | --- | --- |
| ***Self-injury*** |  |  |
| Have you ever harmed or hurt yourself on purpose? | Yes | Receives next question (only included in the DNBC-18) |
|  | No  Do not know | *No self-injury* |
| How often have you harmed or hurt yourself on purpose within the last year? | One time 2-5 times 6-10 times More than 10 times | *Self-injury* |
|  | At no time  Do not know | *No self-injury* |
| ***Suicide ideation*** |  |  |
| Have you ever thought about taking your own life (even though you would not do it)? | Yes | Receives next question (only included in the DNBC-18) |
|  | No  Do not know | *No suicide ideation* |
| How often have you had these thoughts within the last year? | One time 2-5 times 6-10 times More than 10 times | *Suicide ideation* |
|  | At no time  Do not know | *No suicide ideation* |
| ***Suicide attempt*** |  |  |
| Have you ever tried to take your own life? | Yes | Receives next question (only included in the DNBC-18) |
|  | No  Do not know | *No suicide attempt* |
| Have you tried to take your own life within the last year? | Yes | *Suicide attempt* |
|  | No  Do not know | *No suicide attempt* |

**Table S2.** Items on EDs in the DNBC-18 and wave 8 of the COVID-19 survey

| **Questions in the DNBC** | **Response options** | **Used for** |
| --- | --- | --- |
| How much do you weigh? | 150-220 cm | BMI |
| How tall are you? | 30-220 kg | BMI |
| During the past year, how often have you felt too fat? | Never Rarely  Occasionally Often  Every day Do not know | High weight/shape concern |
| During the past year, how often have you felt that you wanted to be thinner? | Never Rarely  Occasionally Often  Every day Do not know | High weight/shape concern |
| During the past year, how often have you been worrying about having too much fat on your body? | Never Rarely  Occasionally Often  Every day Do not know | High weight/shape concern |
| Have you fasted (not eaten anything for at least one day or longer) to lose weight or avoid gaining weight? | Never  Less than once a month  1-3 times a month  Once a week Several times a week  Every day  Do not know | Fasting |
| Have you in the past year exercised to lose or avoid gaining weight? | Never Less than once a month 1-3 times a month Once a week Several times a week Every day Do not know | Excessive exercise |
| Has it been difficult for you to attend school or work because you spend a lot of time exercising? | No  Yes, occasionally Yes, often Do not know | Excessive exercise |
| Do you feel guilty if you skip a workout? | No  Yes, occasionally  Yes, often I never skip a workout | Excessive exercise |
| In the last year, have you been binge eating? | Never Less than once a month  1-3 times a month  Once a week Several times a week  Every day  Do not know | Binge eating |
| Did you feel that you could not stop eating even though you wanted to? | Yes  No Do not know | Binge eating |
| Have you thrown up on purpose to lose weight or avoid gaining weight? | Never Less than once a month  1-3 times a month  Once a week Several times a week  Every day  Do not know | Purging |
| Have you taken laxatives or other medications to lose weight or avoid gaining weight? | Never Less than once a month  1-3 times a month  Once a week Several times a week  Every day  Do not know | Purging |
| Did you eat until you had a stomach-ache or felt sick? | Yes  No Do not know | Abdominal pain |
| Did you feel bad or guilty after eating too much food? | Yes  No Do not know | Guilt |

**Table S3.** Categorasation of threshold and sub-threshold EDs in the DNBC-18 and wave 8 of the COVID-19 survey

| **Threshold ED** |  |
| --- | --- |
| AN | BMI<18.5 AND (high weight/shape concern OR at least monthly fasting OR engaged in excessive exercise) |
| BN | BMI ≥18.5 AND weekly binge eating AND (weekly purging OR engaged in excessive exercise) |
| BED | Weekly binge eating AND experiencing abdominal pain AND guilt AND absence of purging |
| Threshold ED (total) | Threshold AN, BN OR BED |
| **Sub-threshold ED** |  |
| AN | BMI ≥18.5 to <19.5 AND (high weight/shape concern OR at least monthly fasting OR engaged in excessive exercise) |
| BN | BMI≥18.5 AND at least monthly binge eating AND (at least monthly purging OR engaged in excessive exercise) where threshold BN cases are excluded |
| BED | Monthly binge eating AND experiencing abdominal pain AND guilt AND absence of purging |
| PD | Weekly purging AND binge eating absent less than monthly |
| Sub-threshold ED (total) | Sub-threshold AN, BN, BED OR PD |

**Table S4.** Characteristics of baseline population and participants (unweighted)

|  | **Longitudinal setup** | | |  | **Repeated cross-sectional setup** | |
| --- | --- | --- | --- | --- | --- | --- |
| **Variable** | **Baseline**  **N=67,346** | **18-year**  **N=27,441** | **Wave 8**  **N=7,597** |  | **Baseline**  **N=58,638** | **18-year**  **N=24,625** |
|  | **%** | **%** | **%** |  | **%** | **%** |
| **Gender** |  |  |  |  |  |  |
| Male | 51 | 44 | 33 |  | 51 | 44 |
| Female | 49 | 56 | 67 |  | 49 | 56 |
| **Age at wave 8** |  |  |  |  |  |  |
| 19-20 years | .. | .. | 52 |  | .. | .. |
| 21-23 years | .. | .. | 48 |  | .. | .. |
| **Education^a,b^** |  |  |  |  |  |  |
| Low/medium/high education | .. | 85 | 89 |  | .. | 85 |
| Other education | .. | 10 | 7 |  | .. | 10 |
| Not student | .. | 5 | 4 |  | .. | 5 |
| **Housing composition^b^** |  |  |  |  |  |  |
| Living with both parents | .. | 64 | 68 |  | .. | 64 |
| Living with one parent | .. | 28 | 25 |  | .. | 28 |
| Living without parents | .. | 8 | 7 |  | .. | 8 |
| **Socio-occupational status^c^** |  |  |  |  |  |  |
| High educational level | 23 | 26 | 28 |  | 23 | 25 |
| Medium educational level | 31 | 33 | 35 |  | 31 | 34 |
| Skilled worker | 28 | 26 | 23 |  | 28 | 26 |
| Student | 14 | 12 | 11 |  | 14 | 12 |
| Unskilled worker | 2 | 2 | 3 |  | 2 | 2 |
| Outside the labour market | <1 | <1 | <1 |  | <1 | <1 |
| Unclassifiable | <1 | <1 | <1 |  | <1 | <1 |
| **Maternal age at birth** |  |  |  |  |  |  |
| <25 years | 8 | 6 | 5 |  | 8 | 7 |
| 25-29 years | 35 | 36 | 36 |  | 35 | 36 |
| 30-34 years | 39 | 40 | 41 |  | 39 | 39 |
| ≥35 years | 18 | 18 | 18 |  | 18 | 18 |
| **Parity** |  |  |  |  |  |  |
| 0 | 46 | 48 | 51 |  | 46 | 50 |
| 1 | 38 | 37 | 34 |  | 38 | 35 |
| ≥2 | 16 | 15 | 15 |  | 16 | 15 |
| **Maternal smoking during pregnancy** |  |  |  |  |  |  |
| Not smoking | 74 | 76 | 79 |  | 74 | 76 |
| Stopped smoking | 9 | 9 | 8 |  | 9 | 9 |
| 1-10 g daily | 13 | 12 | 10 |  | 13 | 11 |
| <10 g daily | 4 | 3 | 33 |  | 4 | 4 |

^a^ Low/Medium/High education is defined as high school to higher education (> 4 years) and other education is defined as primary school or other education on same level

^b^ Data collected in DNBC-18

^c^ Highest in the household

**Figure S1.** Flowchart of the baseline population in the longitudinal setup from the DNBC-18 and wave 8 in the COVID-19 survey.

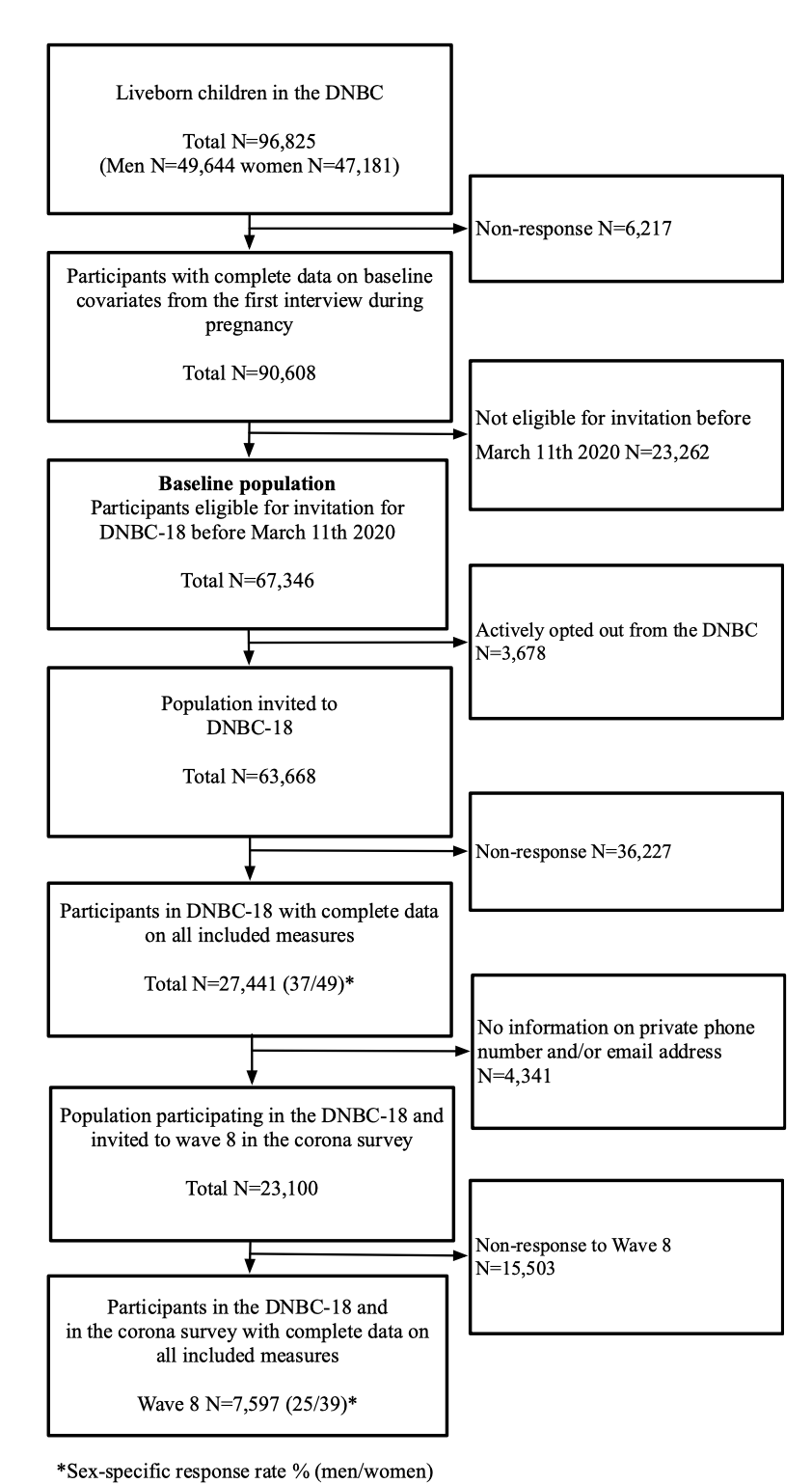

**Figure S2.** Flowchart of the baseline population in the repeated cross-sectional setup from the DNBC-18 in the years 2019-2021.

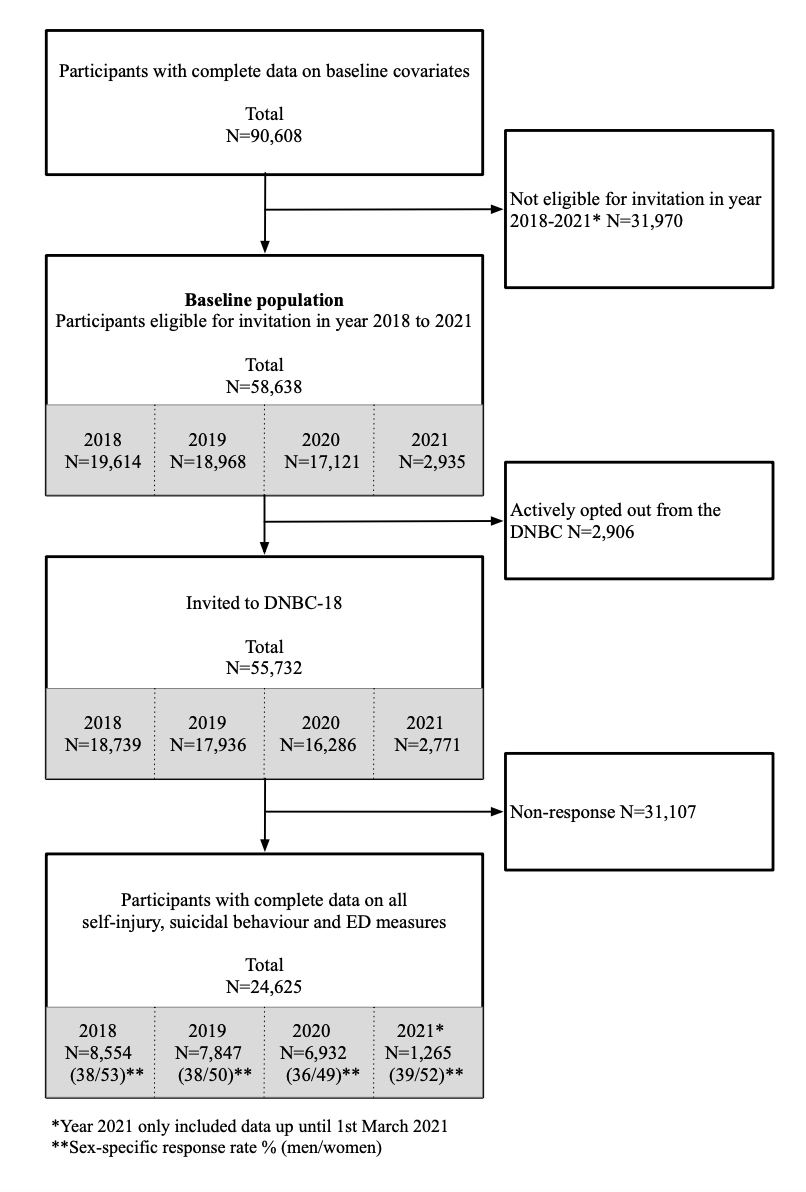

**Figure S3.** Change in the proportion of self-injury, suicide ideation and attempt and EDs from pre to during lockdown in men and women aged 19-20 years when participating in wave 8 based on the longitudinal data collected in the DNBC-18 (N=3,975) and wave 8 of the COVID-19 survey (N=3,975), approximately one year post the initial lockdown.

***a) Self-injury	b) Suicide ideation***

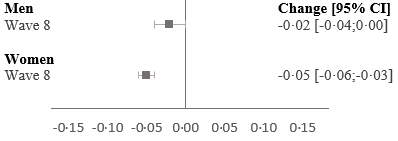

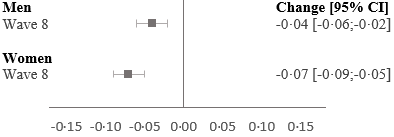

***c) Suicide attempt	d) EDs***

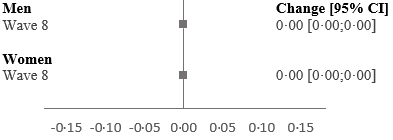

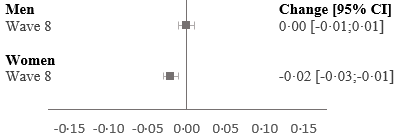

**Figure S4.** Change in the proportion of threshold- and sub-threshold EDs from pre to during lockdown based on a) the longitudinal data collected in the DNBC-18 (N=7,597) and wave 8 of the COVID-19 survey (N=7,597), approximately one year post the initial lockdown and based on b) the repeated cross-sectional setup data collected in the DNBC-18 (N=24,625) in 2018-2021.

***a) Longitudinal data	b) Repeated cross-sectional data***

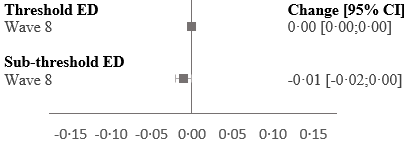

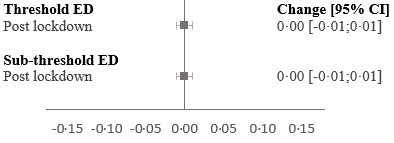
